# Psychedelic mushrooms in the USA: Knowledge, patterns of use and association with health outcomes

**DOI:** 10.1101/2021.09.20.21263824

**Authors:** R Matzopoulos, R Morlock, A Morlock, B Lerer, L Lerer

## Abstract

**Introduction:** Popular media coverage, including of recent positive late stage clinical trials in depression and PTSD, and decriminalization initiatives, are transforming the public perception of psychedelics. However, little is known about levels of knowledge and personal use of psychedelic mushroom(s) (PM) among American adults.

**Methods:** We examined PM use and various measures of health status, quality of life and self-reported mental health outcome measures obtained through a national on-line, cross-sectional survey of adults with a demographic composition representative of the US adult population by region, gender, age, and race (weighted N = 251,297,495) from November 2020–March 2021.

**Results:** General mental health and well-being was a popular reason for PM use (63.6%). PM users were less likely to be overweight than non-users, but overall quality of life (VR-12) was lower for mental health (39.5 vs 45.5). PM users reported significantly higher levels of anxiety (GAD-7 scores of 9.6 vs 5.9) and depression (PHQ-9 scores of 11.2 vs 6.8). They were less likely to have health insurance [OR=0.50 (0.35-0.72)], but reported significantly more healthcare services utilization.

**Discussion and Conclusions:** There is a mismatch between our findings of an association between PM use and poor mental health outcomes, and current discourse on the positive health benefits of PM consumption. A significant number of Americans are already “self medicating” with PM and further research to understand the role of anecdotal knowledge and pseudoscientific information in PM uptake. There is an urgent need for a PM use-related national harm reduction strategy.

## INTRODUCTION

Psychedelics (serotonergic hallucinogens) have powerful and generally predictable psychoactive effects, influencing perception, mood and cognition (1). Following a long hiatus globally linked to restrictive legislative policies, the past decade has seen an exceptional resurgence of research into the therapeutic potential of psychedelics, particularly psilocybin (2), for the treatment of conditions including depression, post-traumatic stress disorder (PTSD) and addiction (3)(4)(5). By December 2020, 70 clinical studies were registered on clinicaltrials.gov, of which 70% were still underway. There is also growing evidence of the potential utility of psychedelics for a range of mental health conditions, pain and neurodegenerative disorders (6)(7)(8)(9).

Biomedical research interest in psychedelics, has been mirrored in a wave of largely positive public discourse (10). In May 2021 the *New York Times* led a front page article heralding a “psychedelic revolution” (11) following publication of extremely promising Phase 3 clinical trial results on MDMA-assisted psychotherapy for PTSD (12). There is also renewed interest in psychedelics in popular culture, from psychedelic content on streaming media such as the Netflix series *The goop Lab* (13) and Vice Media’s *Hamilton’s Pharmacopeia*, and increasing popularity in social media apps such as *Clubhouse*, which posts weekly live podcasts including weekly content such as *The Psychedelic News Hour* (14). A successful psilocybin legalization initiative in Oregon, possible impending psychedelics legalization in California and psychedelic fungi and plant possession decriminalisation ordinances in several cities across the US (including Ann Arbor, Washington DC and Oakland) are driving public policy in the direction of a future recognition of psychedelics as potentially useful for health and wellness. There is however some concern that this wave psychedelics legalization is not being accompanied by evidence-based regulation (15). Investment in psychedelics has increased considerably, with a number of psychedelics biotechnology and services companies listed on the US and Canadian public markets and the launch of the world’s first psychedelics ETF (exchange traded fund) (16).

As compared to opioids, alcohol and tobacco, psychedelics have low addictive potential and benign toxicity profiles (1)(7). There is some evidence users experience reduced psychological distress and suicidality compared to users of other “recreational or illicit” drugs (17). Their perceived safety of psychedelics may be affirmed to some extent by their popular acceptance. Psychedelics are included within the broad category “hallucinogens” in the National Survey on Drug Use and Health (NSDUH), which collects lifetime, past-year and past month drug use estimates representative of persons aged 12 and over in each state and DC. The most recent survey data indicate a significant increase in hallucinogen use from 4.69 million to 6.01 million between 2015 and 2019. The increase was specific to LSD (60% increase) and a 95% increase in other unspecified hallucinogens, a category that included the entheogens (i.e. psychoactive substances historically used for religious or spiritual purposes) peyote, mescaline and psilocybin, whereas PCP and ecstasy use declined over this period (18).

Psilocybin, which is the main psychoactive ingredient in more than 200 species of psychedelic mushrooms (PM) (19), has a particularly benign safety profile and possible positive health effects (20). It has been shown to be comparable to traditional selective serotonin reuptake inhibitors, serotonin-norepinephrine reuptake inhibitors in treating major depressive disorder under clinical trial conditions than (3)(21). Research into psilocybin and pain – a highly prevalent and serious health challenge in the US – is less far advanced, but no less appealing (7). Alongside this recent increase in information about promising therapeutic use the COVID-19 pandemic has imposed considerable societal stress, which adds to the propensity to self-medicate as a coping mechanism. This is manifest in the US with an increase in drinking and alcohol related harms (22), increased anxiety, depression, and social isolation among people with substance use disorders (23), and rising addiction rates associated with stress, personal loss and grief associated with the disease (24).

The purpose of this research was to ascertain knowledge about PM use and therapeutic need among American adults. We compared socio-demographic data for PM users and non-psychedelic users and explored the associations between PM use and various measures of health status, health related quality of life and self-reported mental health outcome measures. We also assessed whether two important contemporary events in the US that may have impacted on mental health, namely the COVID-19 pandemic and the national election and its aftermath, were associated with increased use.

## METHODS

### Data collection

Data were collected through an on-line cross sectional survey of adults (18 years or older) residing in the US in accordance with Acumen Health Research Institute’s (AHRI) established survey procedure. A random stratified sampling framework ensured a community-based sample with a demographic composition representative of the US adult population by region, gender, age, and race, according to the US Census and its standard classifications (US Census American Community Survey 5-year estimate, 2015–2019). Participants were recruited through AHRI’s online research panels that were fielded monthly between November 2020 and March 2021, with each fielding lasting up to 7 days. The survey targeted approximately 1,000-2,000 respondents per month. We applied analysis weights to account for selection probabilities. Multiple quality control processes integrated throughout data collection, included digital finger printing technologies to validate unique respondents and to ensure study data was comprised of non-fraudulent responses. A detailed methodology has been published previously (25).

### Variables

Participants reported on their individual demographic characteristics, educational attainment, health status and knowledge and use of psychedelics in the last 12 months. Estimated participant median household income was derived from US Census American Community Survey 5-year estimate 2015-2019 data for the participant’s zip code. Self-reported co-morbidities were summarised according to the Charlson Comorbidity Index (CCI), a survival stratification tool that assesses the comorbidity risk associated with several conditions and estimates 10-year survival in patients suffering from multiple comorbidities.(26,27) Health-related quality of life (HRQoL) was assessed using the Veterans RAND 12-Item (VR-12) physical component summary (PCS), mental component summary (MCS), and health utility (VR-6D) (28). Anxiety was assessed with the Generalized Anxiety Disorder 7-item (GAD7) (29). Depression was assessed with the Patient Health Questionnaire 9-item (PHQ9) (30). A question about the positive uses of psilocybin “In the last 6 months I heard more than usual about the positive uses of psychedelic drugs (e.g., psychedelic mushrooms) for mental health issues (depression, PTSD, addiction, etc.)?” was rated on a 5-point Likert scale from 0 (strongly disagree) to 4 (strongly agree).

### Analysis

Weighting was carried out according to the census data based on age, sex and region using a Taylor Series Linearization (TSL) method for estimating population characteristics controlling for complex sample survey data. PM users were compared with participants that had not taken a psychedelic in the past 12 months according to population level characteristics, views on psychedelics, treatments, co-morbidities and HRQoL. Users of other psychedelics and combined use of PMs and another psychedelic were excluded from the comparative analysis. An unweighted multivariate logistic regression model controlling for sex, age, race, region, education, employment, CCI score, GAD7, PHQ9 and health insurance predicted psilocybin use. Odds ratios (OR) with 95% confidence intervals are reported.

### Ethics

Respondents were required to be 18 years old or older to participate in the study. Participants confirmed their voluntary agreement to participate and were informed they could leave the survey at any time. Each participant was compensated for their time spent participating according to a reward system based on marketplace points. Participants who fully completed our survey received points worth between $1.40 and $2.45. The actual amount of points awarded to each respondent was determined by the research panel’s incentive policy.

The survey was deemed Institutional Review Board-exempt as all responses were anonymized, aggregated, and could not be related back to individual participants.

## RESULTS

A total of 7,139 participants were included in the sample – a response rate of 83.8%. Psychedelic use in the last year was confirmed by 526 participants (or 7.4% of the total unweighted sample), which included 134 participants (3.8%) who had used PMs and other psychedelics and 122 (3.6%) who had used PMs exclusively. Weighted to the US adult population this represented 7.1% or 17,862,102 adult psychedelic users overall (95% CI: 6.5-7.7), of which 49.2% or 8,780,328 adults (95% CI: 44.7-53.6) were PM users.

We compared the 4,121,102 (1.7%, 95% CI: 1.4-2.1) adults who used only PMs with 233,435,392 (98.3%, 95% CI: 97.9-98.6) adults who did not use any psychedelics. PM users were significantly more likely to be male (68.3% [CI: 59.3-76.1] compared to 47.2% [CI 45.9-48.5] among non-psychedelic users), younger (mean age 38.1 vs 48.6 years), Hispanic/Latino (16.5% vs 8.1%) and reside in the US’s Western Region (33.8% vs 23.0%). Other demographic and socio-economic measures such as income, employment and educational attainment were similar.

In terms of health status (Table 2), PM users were significantly less likely to be overweight than non-users, but reported significantly higher levels of certain comorbid conditions, namely depression, anxiety, migraines and insomnia. This was affirmed by quality of life measures for anxiety (GAD-7 score), and depression (PHQ-9 score). Overall quality of life (VR-12) was also lower for PM users as measured by lower mental health scores (39.5 vs 45.5) and health utility (0.63 vs 0.69), and higher levels of anxiety (GAD-7 scores of 9.6 vs 5.9) and depression (PHQ-9 scores of 11.2 vs 6.8) than non-users. They were also significantly more likely to report healthcare resource utilization, particularly urgent care (20.8% vs 9.9%) and hospitalization (9.2% vs 3.9%).

**Table 1.**
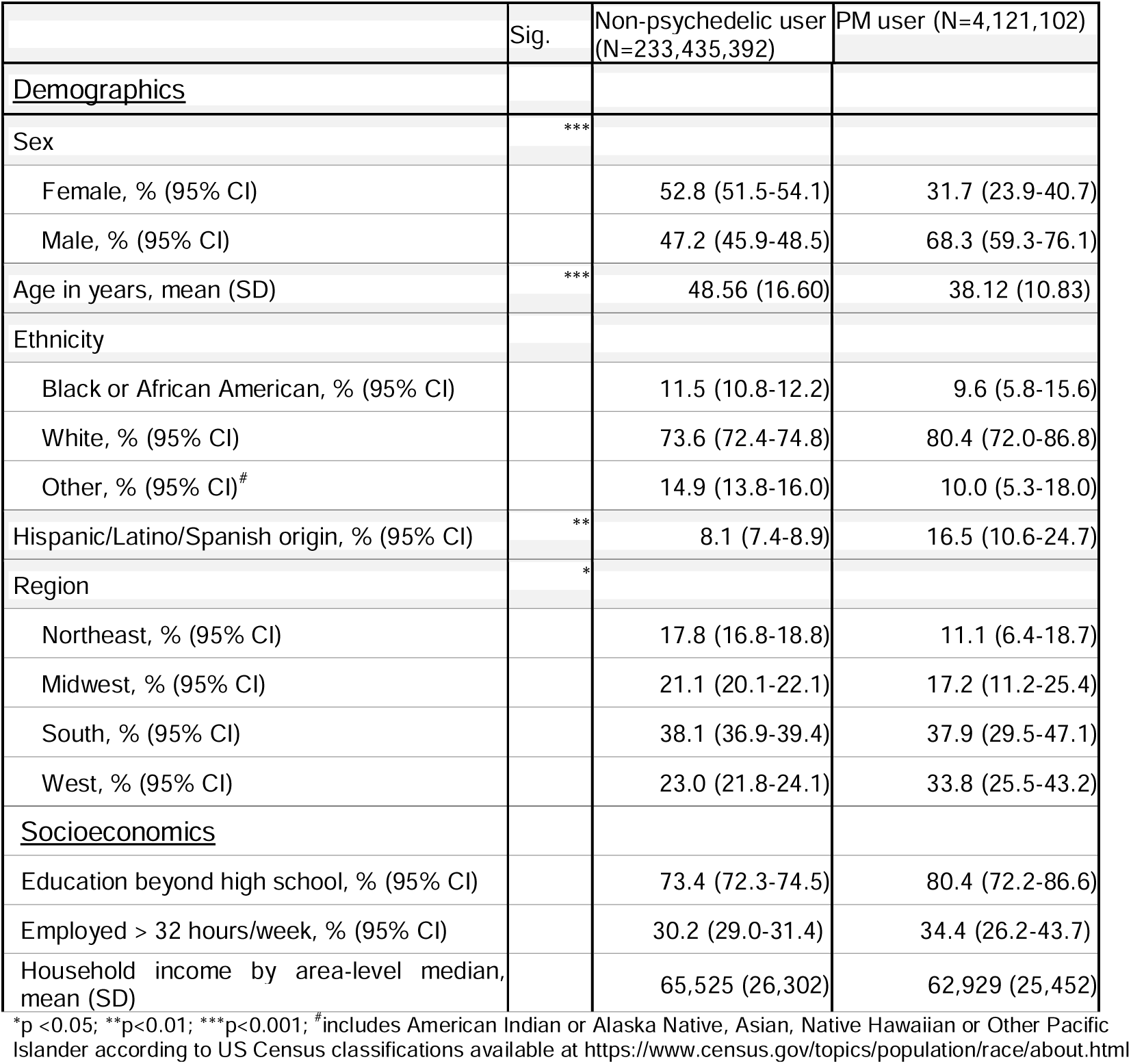
Non-psychedelic user and PM user characteristics, US Adult population 2020/21 (weighted)

|  | Sig. | Non-psychedelic user<br>(N=233,435,392) | PM user (N=4,121,102) |
| --- | --- | --- | --- |
| <u>Demographics</u> |  |  |  |
| Sex | *** |  |  |
| Female, % (95% CI) |  | 52.8 (51.5-54.1) | 31.7 (23.9-40.7) |
| Male, % (95% CI) |  | 47.2 (45.9-48.5) | 68.3 (59.3-76.1) |
| Age in years, mean (SD) | *** | 48.56 (16.60) | 38.12 (10.83) |
| Ethnicity |  |  |  |
| Black or African American, % (95% CI) |  | 11.5 (10.8-12.2) | 9.6 (5.8-15.6) |
| White, % (95% CI) |  | 73.6 (72.4-74.8) | 80.4 (72.0-86.8) |
| Other, % (95% CI)<sup>#</sup> |  | 14.9 (13.8-16.0) | 10.0 (5.3-18.0) |
| Hispanic/Latino/Spanish origin, % (95% CI) | ** | 8.1 (7.4-8.9) | 16.5 (10.6-24.7) |
| Region | * |  |  |
| Northeast, % (95% CI) |  | 17.8 (16.8-18.8) | 11.1 (6.4-18.7) |
| Midwest, % (95% CI) |  | 21.1 (20.1-22.1) | 17.2 (11.2-25.4) |
| South, % (95% CI) |  | 38.1 (36.9-39.4) | 37.9 (29.5-47.1) |
| West, % (95% CI) |  | 23.0 (21.8-24.1) | 33.8 (25.5-43.2) |
| <u>Socioeconomics</u> |  |  |  |
| Education beyond high school, % (95% CI) |  | 73.4 (72.3-74.5) | 80.4 (72.2-86.6) |
| Employed > 32 hours/week, % (95% CI) |  | 30.2 (29.0-31.4) | 34.4 (26.2-43.7) |
| Household income by area-level median, mean (SD) |  | 65,525 (26,302) | 62,929 (25,452) |

\*p < 0.05; \*\*p < 0.01; \*\*\*p < 0.001; <sup>#</sup>includes American Indian or Alaska Native, Asian, Native Hawaiian or Other Pacific Islander according to US Census classifications available at https://www.census.gov/topics/population/race/about.html

**Table 2.** Non-psychedelic user and PM user health, quality of life and health-seeking behaviour, US Adult population 2020/21 (weighted)

|  | Sig. | Non-psychedelic user<br>(N=233,435,392) | PM user<br>(N=4,121,102) |
| --- | --- | --- | --- |
| <u>Health</u> |  |  |  |
| Body mass index (BMI) in lbs/in<sup>2</sup>, mean (SD) | *** | 28.16 (7.11) | 26.01 (5.43) |
| Comorbidities (total reported in past year), mean (SD) |  | 4.07 (3.66) | 4.20 (3.10) |
| Specific comorbid conditions experienced in past year(1) |  |  |  |
| Depression (self-reported), % (95% CI) | *** | 28.3 (27.2-29.5) | 56.8 (47.5-65.7) |
| Anxiety (self-reported), % (95% CI) | *** | 32.8 (31.6-34.0) | 49.1 (40.0-58.3) |
| Chronic pain incl back, neck (self-reported), % (95% CI) |  | 44.5 (43.3-45.8) | 46.7 (37.6-55.9) |
| Allergies (self-reported), % (95% CI) |  | 28.7 (27.6-29.9) | 32.2 (24.3-41.1) |
| Migraines (self-reported), % (95% CI) | *** | 18.6 (17.6-19.6) | 31.2 (23.3-40.5) |
| Insomnia (self-reported), % (95% CI) | * | 20.6 (19.6-21.7) | 30.0 (22.2-39.0) |
| Hypertension (self-reported), % (95% CI) |  | 23.6 (22.5-24.7) | 16.8 (11.0-24.7) |
| Diarrhea, chronic, or more than occasional (self-reported), % (95% CI) |  | 14.2 (13.3-15.1) | 10.7 (6.3-17.5) |
| Sleep apnea (self-reported), % (95% CI) |  | 8.2 (7.5-8.9) | 10.1 (5.8-17.1) |
| GERD (self-reported), % (95% CI) |  | 13.6 (12.7-14.5) | 9.7 (5.3-16.9) |
| Constipation, chronic, or more than occasional (self-reported), % (95% CI) |  | 13.7 (12.8-14.6) | 8.9 (5.0-15.3) |
| Hyperlipidemia (self-reported), % (95% CI) |  | 15.5 (14.6-16.5) | 8.7 (4.6-15.6) |
| CCI score, mean (SD) |  | 0.48 (1.04) | 0.60 (0.98) |
| Anxiety (GAD-7 score), mean (SD) | *** | 5.88 (5.79) | 9.62 (5.76) |
| Anxiety (GAD-7 ≥ 10), % (95 CI) | *** | 25.3 (24.2-26.4) | 50.2 (41.0-59.3) |
| Depression (PHQ-9 score), mean (SD) | *** | 6.75 (6.79) | 11.23 (6.99) |
| Depression (PHQ-9 ≥ 10), % (95 CI) | *** | 29.5 (28.4-30.7) | 56.1 (46.6-65.0) |
| <u>Quality of life</u> |  |  |  |
| VR-12 |  |  |  |
| MCS, mean (SD) (t-score) | *** | 45.53 (12.44) | 39.49 (11.25) |
| PCS, mean (SD) (t-score) | * | 45.28 (10.45) | 43.51 (9.24) |
| Health utility (VR-6D), mean (SD) | *** | 0.69 (0.12) | 0.63 (0.10) |
| <u>Health-seeking behaviour</u> |  |  |  |
| Primary care physician visit, % (95% CI) |  | 52.4 (51.1-53.7) | 51.1 (41.9-60.3) |
| Specialist, % (95% CI) |  | 28.6 (27.5-29.8) | 23.2 (16.4-31.7) |
| Other healthcare provider, % (95% CI) | * | 19.7 (18.7-20.7) | 28.2 (20.7-37.1) |
| Urgent care, % (95% CI) | *** | 9.9 (9.2-10.7) | 20.8 (14.3-29.3) |
| Outpatient procedure or surgery, % (95% CI) |  | 8.6 (7.9-9.3) | 7.2 (3.5-14.2) |
| Emergency department, % (95% CI) |  | 10.1 (9.4-10.9) | 12.9 (8.1-20.1) |
| Hospitalization, % (95% CI) | ** | 3.9 (3.4-4.4) | 9.2 (5.2-15.8) |

\* p <0.05; \*\* p<0.01; \*\*\* p≤0.001
(1) Co-morbid conditions are shown for the 10 most reported conditions for each cohort.

PM users consistently and significantly held more favorable views of the positive potential for PM use for the treatment of a range of conditions than non-users (Table 3), including for general mental health, well-being and personal development (65.8% vs 15.8%), and managing diagnosed (50.9% vs 16.6%) and self-diagnosed psychiatric conditions (34.7% vs 7.6%). This was consistent with their rationale for using PMs, with general mental health and well-being being the most common (63.6%), followed by use for diagnosed psychiatric conditions (31.8%) and self-diagnosed conditions (19.0%). PM users were also significantly more likely than non-users to have heard more frequent positive reporting of the use of psychedelic drugs for mental health issues (depression, PTSD, addiction, etc.) in the last six months than previously (mean score 2.06 vs 3.56 out of a scale of 1 “strongly agree” to 5 “strongly disagree”). Frequency of use did not seem to have been influenced on account of either COVID-19 or election politics. PM use was significantly associated with medical treatment following use (17.9% of users).

**Table 3.** Non-psychedelic user and PM user, knowledge and use of PMs for health and wellbeing, US Adult population 2020/21 (weighted)

|  | Sig. | Non-psychedelic user<br>(N=233,435,392) | PM user<br>(N=4,121,102) |
| --- | --- | --- | --- |
| <u>Health and wellbeing knowledge</u> |  |  |  |
| More frequent positive information about PM use for mental health in last 6 months (score)(range:1, strongly agree to 5, strongly disagree), mean (SD) | *** | 3.56 (1.20) | 2.06 (1.07) |
| Positive potential for general mental health, well-being and personal development (preventive), % (95% CI) | *** | 15.8 (14.9-16.7) | 65.8 (56.7-73.9) |
| Positive potential managing a diagnosed psychiatric condition – PTSD, depression, addiction, etc (curative), % (95% CI) | *** | 16.6 (15.7-17.6) | 50.9 (41.7-60.1) |
| Positive potential managing a self-diagnosed condition/concern – relationship issue, bereavement, trauma, addiction, etc (curative), % (95% CI) | *** | 7.6 (7.0-8.3) | 34.7 (26.5-44.0) |
| No knowledge, % (95% CI) | *** | 69.7 (68.8-70.9) | 13.6 (8.3-21.4) |
| <u>Reasons for use</u> |  |  |  |
| General mental health, well-being and personal development (preventive), % (95% CI) | – | – | 63.6(54.2-72.0) |
| Managing a diagnosed psychiatric condition – PTSD, depression, addiction, etc (curative), % (95% CI) | – | – | 31.8 (23.9-41.0) |
| Managing a self-diagnosed condition/concern – relationship issue, bereavement, trauma, addiction, etc (curative), % (95% CI) # | – | – | 19.0 (13.0-27.0) |
| <u>Frequency of use</u> |  |  |  |
| Increased on account of COVID-19 | – | – | 14.7 (9.4-22.3) |
| Decreased on account of COVID-19 | – | – | 17.9 (11.9-25.9) |
| No impact | – | – | 66.6 (57.5-74.7) |
| Increased on account of election politics | – | – | 13.1 (8.1-20.5) |
| Decreased on account of election politics | - | - | 11.6 (7.0-18.7) |
| No impact | - | - | 74.3 (65.5-81.5) |
| <u>Potential harm</u> |  |  |  |
| Seeking medical treatment following use | - | - | 17.9 (11.9-26.0) |

\* p <0.05; \*\* p<0.01; \*\*\* p<0.001

The multivariate logistic regression analysis explored the correlation between various factors and PM use (Table 4). Factors predictive of PM use included being male [OR=1.53 (1.09-2.15)] and reporting worse health [OR=1.42 (1.22-1.65)]. Those with health insurance [OR=0.50 (0.35-0.72)], increased age [OR=0.92 (0.90-0.93)], and relative to those living in the west US census region, those living in the northeast [OR=0.27 (0.15-0.50)], midwest [OR=0.34 (0.20-0.56)], and south [OR=0.38 (0.26-0.55)] were less likely to report PM use.

**Figure 1.**
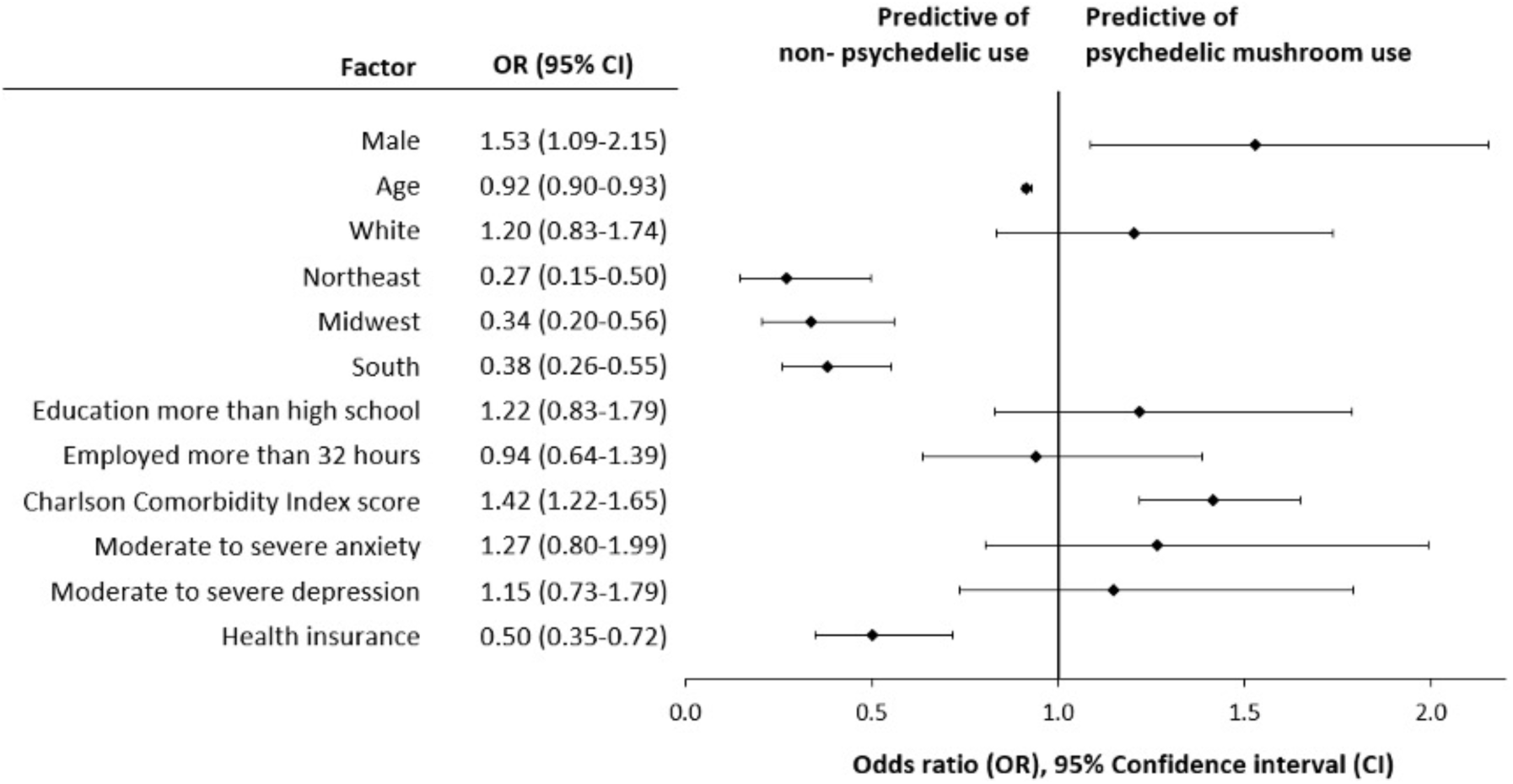
Results of multivariate logistic regression model predicting past year psychedelic mushroom use for selected demographic and educational factors and health and comorbidity indicators.

## DISCUSSION

The objective of the present analysis was to ascertain knowledge about PMs among American adults and explore associations between PM use and various general and self-reported mental health outcome measures. Our findings confirm the popularity of psychedelics more broadly and PMs specifically among the US adult population (19,31,32). Estimated past-year psychedelic use of 7,7%, which equated to approximately 17,9 million adult Americans (95%CI: 16.4 – 19,4 million) was almost three times higher than recorded in the NSDUH, which estimated past-year hallucinogen use at just more than 6 million persons in 2019. Similarly estimated past-year PM use of 4.1 million adult Americans (95%CI: 3.4 – 5.1 million) was also significantly higher than the 2019 NSDUH, which reported past year use of *all hallucinogens other than LSD, PCP and ecstasy* at less than 1 million persons (18).

The popularity of PMs amongst psychedelic users, approximately half of all past year psychedelic use in our study, was not unexpected. Psilocybin has a relatively benign reputation among psychedelics generally, which is further enhanced by PMs being a natural rather than synthetic source of psilocybin. Among the different classes of psychedelics, classic tryptamines, which include psilocybin, are uniquely associated with a decreased likelihood of psychological distress and suicidal thinking and are widely considered to hold the greatest therapeutic potential among lifetime users(33). This was confirmed by PM users identified in our study. Despite this favourable view we found concerning associations between of PM use and negative mental health outcomes including anxiety, depression, comorbidities and medical treatment following use. Although associations for anxiety and depression were not significantly associated with PM use in the logistic regression analysis, PM users were still significantly associated with comorbidities and were less likely to have health insurance. Higher utilization of healthcare resources could increase costs and, in turn, drive a gap in care for PM users.

These findings were distinctly different from a recent international study which showed that regular users of psychedelic drugs had less psychological stress compared to occasional users and non-users, suggesting that the use of psychedelics might either be a protective factor or that people with certain traits were more prone to frequently use psychedelic drugs (34). This difference could either be ascribed to our study focusing on any use rather than regular use, or that PM users in the US comprise a distinctly different demographic grouping than their international counterparts.

Either way, more certainty is needed about dosage and treatment effect windows for PMs, as well as the potential addictive effects and side effects of consistent use arising from interactions of psilocybin with dopamine and dopamine receptors (35), as psilocybin stimulates dopamine receptors indirectly (1). Ongoing litigation of pharmaceutical companies implicated in the opioid epidemic in the US (36) emphasises the importance of a considered approach in developing a comprehensive understanding of risks and benefits not only to individual users, but also for their families and communities.

While use of PMs may have been expected to increase during the COVID-19, as individuals seek to self-medicate to deal with the trauma of isolation and anxiety about the virus (37), interest is unlikely to wane in its aftermath. COVID-19 has thrown mental health disorders into sharp focus and therapeutic interventions are likely to find renewed impetus. Almost one-third of COVID-19 cases with serious complications may experience PTSD (38) while the full impact on grieving families and communities will only become apparent in the future. However, the overwhelmingly positive coverage of psychedelics requires some circumspection and some of the correlates of LSD use serve as a warning. These include higher levels of antisocial behavior as measured through interaction with the criminal justice system and the presence of comorbid mental health and substance abuse disorders. (39).

Classical serotonergic psychedelics do not cause neurotoxic effects, organ damage or long-term neuropsychological deficits (1). Acute psychological effects remain a possibility, but these can be averted through supervised administration of perception-altering doses and there are also distinct medicinal uses of sub-perceptual doses or molecules lacking perceptual effects (40). There is also limited possibility of physical dependence. Animal studies have shown that they lack reinforcing properties that induce self-administration, which limits the potential for abuse as well as withdrawal symptoms (41,42). This is consistent with a US population-based study that indicated no association between lifetime use of psychedelics generally, or specific psychedelics including psilocybin, and increased rate of mental illness and prolonged, or re-occurring, perceptual effects(43).

Less information is available for novel psychedelics, which are becoming increasingly popular and seem less benign. Sexton et al. (2019) found a greater likelihood of past year suicidal thinking and past year suicidal planning among lifetime users of novel psychedelics (44). Subsequent analysis of the NSDUH found that novel phenethylamine specifically was associated with increased suicidal thinking and suicidal planning (33).

Across cultures, people try to make sense of the world around them, including how likely it is that a negative outcome will occur for them and what will increase or reduce that risk - *lay epidemiology* (45). Their decisions are complicated not only by the expanding variety of available psychedelics, but also their potential application for an increasing range of therapeutic uses. It is known, for example, that psychedelics exert significant modulatory effects on immune responses by altering signalling pathways involved in inflammation, cellular proliferation, and cell survival (46). Preclinical research has shown that psychedelics promote structural and functional neural plasticity in key brain regions linked to psychological functioning (9). A recent milestone publication provides the scientific hypothesis for low doses of psychedelics exerting effects on mental well-being and neurological healing through indirect modulation of the gut-brain axis (47). Consequently, therapies involving psychedelics are among the innovative treatments for traumatic brain injury (48).

Population-based research such as the present study could help to indicate receptivity for possible therapeutic application. In addition it can be argued that there is a need for such data to supplement traditional confirmatory with real world evidence and studies that can inform pragmatic trial designs (3). For example the recently reported association between lifetime classic psychedelic use and cardiometabolic diseases (32) would be strengthened if the association was found to be consistent when measured against more recent use.

Our study design presented both strengths and limitations. The use of an online data collection platform reduced the risk of stigma by ensuring privacy and confidentiality, which may explain the considerably higher past year estimates than were reported in other surveys. Although we made every effort to avoid sampling biases and to ensure the validity of information, standard survey limitations arising from self-reporting still applied and may have been amplified by including only respondents who had access to a computer and were willing to take an online survey. Another limitation was that comparisons were made with no alpha adjustment resulting in an increased risk of Type 1 errors. We excluded users of other psychedelics and users that combined PM use with use of other psychedelic to avoid confounding, but in so doing may have missed other significant associations. The inability to distinguish between different users may have introduced some selection bias – people using psychedelics parsimoniously for the first time were more likely to be included, whereas experienced “psychonauts” using a wide range of designer psychedelics could have been excluded (49). We also note that the timing of the study was unusual, in that it was conducted at the height of the COVID-19 pandemic, which may make the findings less generalisable to surrounding time periods. In its favour, questions about recent rather than lifetime PM use present information that is current and arguably more relevant for a public health response.

## CONCLUSIONS

The present research clearly demonstrates the utility of population-based understanding of entheogen use, among which PMs remain particularly popular. Despite the overwhelmingly positive reputation of life time use of PMs conferring protective qualities our study has shown a disturbing association with negative physiological and mental health outcomes. The extent to which uptake is influenced by emerging scientific evidence versus anecdotal or pseudoscientific knowledge remains unclear, but is incidental to the need for the development of guidelines to optimize therapeutic use. Future research should include regularized application of our population-level methodology surveys to assess changes over time as well as additional modules to identify key drivers of use and misuse for different entheogens, and to indicate possible therapeutic applications for specific conditions. Separate analysis of other psychedelic use, exclusively and in combination, should also be considered.

## Data Availability

Data will be made available prior to publication.

## Author Contributions

Richard M was the primary author who drafted the manuscript summarizing the findings. Rob M and AM informed methodology, conducted analyses and were responsible for ensuring analyses were interpreted correctly. LL, corresponding author, conceptualised the study and, along with BL, contributed meaningful pharmacological expertise to aid in the interpretation of results. All authors contributed to and approved of the final version of the manuscript.

## Funding

BYAS sponsored Back of the Yards Algae Sciences, Chicago-based sustainable biotechnology company.

## Notes

### Competing Interest Statement

Bernard Lerer and Leonard Lerer hold equity in Back of the Yard Algae Sciences and Parow Entheobiosciences. There are no other conflicts relevant to this work.

### Funding Statement

No external funding was received for this research.

### Author Declarations

Sterling IRB in Atlanta, Georgia, reviewed the protocol (ID: 8249-RJMorlock)and determined that the study was exempt from IRB review pursuant to the terms of the U.S. Department of Health and Human Service's Policy for Protection of Human Research Subjects at 45 C.F.R. S46.104(d) and that the following exemption category applied: Category 2 Exemption (DHHS).

